# Language and cognitive outcomes after pediatric ischemic stroke: The role of linguistic diversity and age at stroke onset

**DOI:** 10.64898/2025.12.09.25341917

**Authors:** Kai Ian Leung, Robyn Westmacott, Nomazulu Dlamini, Elizabeth Rochon, Monika Molnar

## Abstract

Pediatric stroke disrupts ongoing neurodevelopment and often results in long-term cognitive challenges. For bilingual children navigating healthcare systems that underserve linguistically diverse families, these impacts raise critical health equity considerations. While bilingualism has historically been framed as a burden, research suggests it may support development in clinical populations. This cross-sectional study investigated how linguistic diversity relates to language and cognitive outcomes following arterial ischemic stroke, examining moderation by age at stroke onset. Twenty-nine children completed language and executive function assessments; caregivers reported language exposure and socioeconomic information. Analyses showed no main effect of linguistic diversity, but significant interactions with age at stroke. In the neonatal group, greater linguistic diversity was associated with better expressive language, whereas the presumed perinatal group showed the inverse pattern; childhood-onset stroke showed minimal associations. Findings suggest that bilingual environments do not impede, and may support, language following stroke, underscoring the importance of culturally/linguistically responsive care.

## Introduction

Although less common than in adults, pediatric stroke has an estimated incidence of 2–38 per 100,000 children annually and often results in long-term rehabilitation needs and reduced quality of life.^1–3^ As stroke coincides with critical processes during neurodevelopment,^4,5^ impairments may emerge later or manifest as disruptions to developmental milestones.^6,7^ Post-injury, children may experience persistent deficits across multiple domains, including language, cognition and psychosocial functioning,^8–10^ with about half of all stroke patients receiving a learning and/or psychological diagnosis.^11^

Pediatric stroke is heterogeneous in both etiology and presentation, with different profiles potentially presenting with different outcomes reflecting the complex interplay of neurological, environmental and developmental factors.^12,13^ Among neurological determinants, *age at stroke onset* is a well-recognized predictor of post-stroke outcome. Strokes occurring in the presumed perinatal (i.e., acquired perinatally but diagnosed after the neonatal period when neurological signs emerge), neonatal (i.e., symptomatic and occurring within the first 28 postnatal days), and childhood (i.e., symptomatic and occurring between >28 days – 18 years) periods differ in clinical presentation and outcomes.^12^ In cohort studies and reviews, functional post-stroke outcomes vary by age at onset, revealing a complex, non-linear effect. Some studies report comparatively worse cognitive and linguistic outcomes in neonatal vs childhood stroke, other studies show the reverse, and some have reported a U-shaped pattern; this variety highlights that age effects may depend on other lesion characteristics and a mix of profiles (e.g., acute neonatal vs. presumed perinatal cases).^14–17^ Longitudinal studies have also linked earlier stroke onset with specific cognitive risks (independent of other factors), reinforcing age at onset as a critical covariate in pediatric stroke outcome models.^14,18^ Other neurological contributors to post-stroke outcomes also include lesion location, laterality and volume, time since stroke, stroke type, neurologic impairment, and seizure history^17,19–21^

Beyond neurological factors, *environmental influences* also merit greater consideration, particularly within a social determinants of health framework.^22–25^ Structural inequities tied to socioeconomic status (SES) and access to linguistically supportive environments are central health equity considerations, as they may shape developmental trajectories following pediatric brain injury.^18,26,27^ Children from lower SES backgrounds exhibit significantly worse long-term neurologic, cognitive and linguistic outcomes compared to peers from higher SES families.^24,26,28,29^ SES can account for substantial variance (up to 42%) in post-stroke cognitive outcome, at times exceeding the effects of clinical and lesion characteristics.^28^ These patterns highlight that the socioeconomic context is a key environmental determinant of post-stroke outcomes. Yet families from linguistically diverse backgrounds, who may also navigate intersecting socioeconomic or other structural inequities, remain underrepresented in pediatric stroke research.

Nearly half the global population grows up with more than one language.^30^ Assuming stroke incidence is distributed across linguistic groups, roughly half of children who sustain a pediatric stroke come from homes where multiple languages are spoken. Children’s language environments exist on a continuum—from monolingual households where a single language is used exclusively, to bilingual (or multilingual) households where children are exposed to two (or more) languages with varying levels of proficiency, frequency, and context of use (e.g., school, home, other). In this study, we refer to this variation in bi- and multilingual language exposure as linguistic diversity (hereafter, bilingualism is used inclusively to also encompass multilingualism).

Historically, bilingualism was perceived as a burden,^31,32^ with concerns revolving around the idea that managing two languages could delay acquisition or hinder development, especially in neurodivergent individuals.^33,34^ Contemporary research has challenged these assumptions, instead highlighting potential cognitive advantages associated with bilingualism in neurotypical populations. However, this is not without debate.^35,36^ In neurotypical children, bilingualism has been associated with enhanced executive functions, such as attentional control, cognitive flexibility, and working memory.^37^ These advantages are hypothesized to arise from the continuous need to monitor, select, and inhibit competing linguistic systems,^38^ which is said to afford bilinguals automaticity and efficiency in non-linguistic task processing thanks to a modified attentional system.^39^

Within clinical populations, however, evidence remains more limited and mixed with regards to bilingualism effects. Much of the bilingualism literature has focused on populations with pronounced developmental disorders associated with language and cognitive impairments, such as developmental language disorders, autism spectrum disorder and other neurodevelopmental disorders.^40^ These studies have found that bilingual children often perform as well as or better than monolingual peers in language and cognitive outcomes.^41,42^ While some early perspectives suggested that bilingual exposure among children with diverse neurodevelopmental profiles might hinder development through added cognitive demands, emerging studies including Leung et al. indicate that bilingual children may fare as well as, or better than, their monolingual peers on post-stroke language outcomes.^18^ However, questions remain regarding the domain (i.e., cognition vs. language) and task-specificity (i.e., different effects depending on the task within a domain) of these effects and the conditions under which bilingualism may act as a protective factor. Critical variables such as age at stroke, age of second language acquisition, and proportion of bilingual exposure may moderate outcomes, highlighting the need for systematic investigation.

While stroke guidelines recommend comprehensive neuropsychological assessment to document cognitive and language deficits and guide therapy and educational planning,^13^ these guidelines rarely address the needs of bilingual children or families with limited societal language proficiency, leaving clinicians and educators with limited evidence-based direction. Despite the prevalence of bilingualism worldwide, research addressing these issues in pediatric stroke populations is scarce. There is a critical need for evidence that integrates neurological determinants with linguistic and environmental contexts to better understand stroke outcomes in children from diverse language backgrounds.

### Current Study

This study investigates how linguistic diversity influences linguistic (e.g., expressive language, metalinguistic skills) and cognitive outcomes (e.g., executive function, nonverbal reasoning) in children with arterial ischemic stroke (AIS). In evaluating key determinants such as age at stroke and quantity of language exposure, we aim to determine whether bilingualism moderates these developmental outcomes independent of SES effects. This approach strengthens the health-equity evidence base, specifically considering paediatric stroke. In line with recent methodological recommendations in bilingualism research, linguistic diversity was characterized both continuously (proportional indices of language exposure) and categorically (monolingual vs. bilingual groups based on age of acquisition thresholds).^43^ This approach enables examination of both between-group differences and within-group variability in language experience, providing a more nuanced understanding of bilingual effects. To contextualize performance, outcomes were expressed as percentile ranks relative to peers within this sample, rather than external population norms. Analytically, we employed regression-based modeling approaches to test the joint effects of linguistic diversity and biopsychosocial factors on developmental outcomes. These models are well suited for examining both categorical and continuous predictors and for identifying potential moderation by age at stroke onset.

### Study Aims

This study evaluates the impact of linguistic diversity on language and cognitive outcomes in children with AIS, examining whether age at stroke onset moderates these relationships while also adjusting for SES. Building on prior findings from Leung et al.,^18^ which identified language advantages associated with greater linguistic diversity - particularly among children with early stroke onset - this study hypothesizes that linguistic diversity may differentially influence outcomes depending on the age at which the stroke occurred. Given prior moderation by age at stroke, we anticipate differential effects of language exposure across age at stroke onset groups (neonatal, presumed perinatal, childhood). We also expect greater linguistic diversity to be non-detrimental and potentially advantageous for language. Analyses involving the presumed perinatal stroke group are considered exploratory, due to both the diagnostic ambiguity in classifying stroke timing and the absence of a comparable subgroup in the Leung et al. study.^18^

## Materials and Methods

### Participants

Participants were identified through a review of The Hospital for Sick Children (SickKids)’s outpatient clinic charts to screen for a history of pediatric arterial ischemic stroke (AIS). Medical records were examined against predefined eligibility criteria. Children and adolescents (0-18 years at stroke onset) with an ischemic infarct, including single or recurrent events, who were ≥ 6 months post-stroke and whose caregivers and themselves demonstrated sufficient English proficiency for study completion were considered eligible for the study. Exclusion criteria included an inability to complete study procedures (e.g., due to severe intellectual disability, insufficient English proficiency).

### Ethical considerations

Ethical approval for this study was obtained from the SickKids Research Ethics Board (REB# 1000081176). Prior to participation, researchers explained the study objectives and procedures to families as part of the informed consent process. Written informed consent from caregivers and assent from youth was obtained prior to participation.

### Procedure

Eligible families were recruited during routine initial or follow-up neuropsychological assessment appointments at SickKids (Toronto, Ontario). During visits, participants completed neuropsychological testing while caregivers completed questionnaires, requiring approximately one hour. All assessments followed standardized administration procedures, were administered in English and interpreted by the study team comprising a pediatric neuropsychologist, a psychometrist, and supervised doctoral students.

### Measures

To assess language and cognitive functioning, participants completed a battery of standardized, norm-referenced measures (Table 1). Test compatibility with the pediatric stroke population was carefully considered following current recommendations.^44^ Domains of expressive language, metalinguistic awareness and cognition (executive function; response inhibition, switching, monitoring) that have previously been used with neurotypical and neurodivergent bilinguals were targeted. In general, higher scores indicated better performance.

**Table 1.** Domain, neurocognitive skills and associated measures in child participants.

| **Domain** | **Neurocognitive skill** | **Standardized measures** |
| --- | --- | --- |
| Language | Expressive language | CELF-5, Formulated Sentences |
|  | Metalinguistic awareness | CELF-5 Metalinguistics, Making Inferences; Multiple Meanings |
| Cognition | Executive function | D-KEFS - Trail Making Test, Number-Letter Switching<br>D-KEFS - Color-Word Interference Test, Inhibition; Inhibition / Switching |
|  | Nonverbal reasoning | Wechsler (WISC-V / WAIS-IV), Matrix Reasoning |

*Abbreviations:* CELF-5 = Clinical Evaluation of Language Fundamentals – Fifth Edition; D-KEFS = Delis-Kaplan Executive Function System; WISC-V = Wechsler Intelligence Scale for Children – Fifth Edition; WAIS-IV = Wechsler Adult Intelligence Scale – Fourth Edition.

#### Standardized Assessments

##### Clinical Evaluation of Language Fundamentals – Fifth Edition

(CELF-5). The CELF-5^45^ is a gold-standard, language assessment validated for children aged 5-21. The Formulated sentences test evaluated children’s expressive language. During this test, the child formulates a sentence using an orally presented target word(s) with a stimulus picture as reference.

##### Clinical Evaluation of Language Fundamentals – Fifth Edition Metalinguistics

(CELF-5 M). The CELF-5 M^46^ was designed to identify students aged 9-21 who have not acquired the expected levels of communicative competence and metalinguistic ability for their age. The Making Inferences and Multiple Meanings subtests evaluated the child’s ability to identify logical inferences given causal relationships or event chains and interpret different meanings of word and sentence level ambiguities, respectively. In the Making Inferences subtest, the child listens to the examiner describe a situation by its beginning and ending. The child identifies the best two out of four reasons given for the ending and provides an additional reason outside of the list. In the Multiple Meanings subtest, the tester presents a sentence that contained ambiguity at either the word or sentence level. The child is asked to describe two meanings for each sentence that is presented.

##### Matrix Reasoning (MR) subtest of the Wechsler Scales

(Wechsler Intelligence Scale for Children – Fifth Edition [WISC-V] and Wechsler Adult Intelligence Scale – Fourth Edition [WAIS-IV]). MR is a nonverbal test of cognitive ability evaluating abstract reasoning and problem solving, normed for ages 6-16 for the WISC-V^47^ and 16 to 90 for the WAIS-IV^48^. In this test, the child is asked to look at a series of visual patterns with a missing piece and choose the option that correctly completes the pattern.

##### Delis-Kaplan Executive Function System

(D-KEFS). The D-KEFS^49^ is a standardized assessment of higher-level cognition in those aged 8-89 and was used to assess key components of the child’s executive functions. The Color-Word Interference and Trail Making subtests were used. The Color-Word Interference Test measures ability to inhibit a dominant and automatic verbal response. In this test, the child is asked to name colors and read color words, and in some conditions to inhibit reading the word and instead name the ink color, as quickly and accurately as possible. The Trail Making Test measures flexibility of thinking on a visual-motor sequencing task. In this test, the child is asked to connect circles containing numbers or letters in the correct order, and in some conditions to alternate between numbers and letters, as quickly and accurately as possible.

#### Caregiver-Reported Questionnaires

##### Demographics and History Questionnaire

This provided information regarding the participants’ developmental, medical, and family history. Socioeconomic status was operationalized as a composite of household income, caregiver highest education and material deprivation index of the Ontario-Marginalization index (ON-Marg) – a validated index derived from Canadian census data that provides neighborhood-level estimates of material deprivation. The ON-Marg has been operationalized in previous investigations of post-stroke outcomes in adults^50^ and in children.^27^ Other relevant determinants of health (e.g., race/ethnicity, sex) were also recorded.

##### Language Experience and Proficiency Questionnaire

(LEAP-Q). The LEAP-Q^51^ is a reliable and valid tool used to assess language profiles of multilingual children for research use. Linguistic variables, such as current language exposure (e.g., English, other languages), age of acquisition, proficiency and context of use are essential to understanding the patient’s language case history as these are also known independent predictors of performance on language measures. For the purposes of this study, we operationalized linguistic diversity as the proportion of exposure to English versus other language(s) in children’s daily communicative contexts, derived from parent report. Higher linguistic diversity indicates greater exposure to non-English languages (e.g., 50% English, 50% other language(s)), while lower linguistic diversity indicates English-dominant or monolingual English environments (e.g., 100% English). The sample was also categorized as monolingual or simultaneous bilingual (exposed to both languages before age 3), using the same criteria as detailed in Leung et al.^18^

For sample description, we also employed the following parent-rated questionnaires. *Behavioural Rating Inventory of Executive Function – Second Edition* (BRIEF-2)^52^ was used to assess parent-reported impairment of executive functioning skills and self-regulation in everyday environments. The *Child Trauma Screen* (CTS)^53^ was used to screen for children’s exposure to potentially traumatic events that could impact emotional and behavioral functioning (e.g., immigration related trauma, medical trauma). The *Pediatric Symptom Checklist-17* (PSC-17)^54^ was used as a brief psychosocial screening to facilitate recognition of cognitive, emotional, and behavioural impairment.

#### Medical Record Abstraction

Additional clinical data were abstracted from participants’ medical charts. This information was used to characterize the sample and serve as covariates in analyses of language and cognitive outcomes. Data included stroke-specific factors (e.g., age at stroke, lesion location, laterality, time since stroke), neurological status, and relevant medical and developmental history. The latest available Pediatric Stroke Outcome Measure (PSOM)^55^ was used to quantify neurologic outcome across domains.

### Statistical analysis

Primary outcomes were derived from standardized scores from language (CELF-5 Formulated Sentences; CELF-5 M Making Inferences, Multiple Meanings) and cognitive (D-KEFS Number–Letter Switching, Inhibition, Inhibition/Switching; Wechsler Matrix Reasoning) measures. For tasks where non-attempts reflected examiner judgment that the test would be too difficult for the participant, we applied a worst-rank strategy: these cases were assigned the lowest observed rank plus one, ensuring placement at the bottom of the distribution. Scores were then transformed within-sample percentile ranks (0–1, higher = better) computed from cohort ranks 0.5 /. This common scaling enabled cross-task comparisons and mitigated floor effects. This approach was particularly appropriate given the high proportion of bilingual participants, who are usually underrepresented in published norms, and aligns with the ordinal logic of worst-rank imputation. Percentiles reflect relative standing within the study sample rather than population-normative benchmarks. For outcomes with minimal missingness (e.g., Wechsler Matrix Reasoning), analyses were conducted using the native standardized scales.

For each outcome, we fit a linear model: outcome ∼ percentage English exposure (z-scored) × age at stroke (neonatal/childhood/presumed) + composite SES (z-scored). To minimize overfitting in N≈25–29, clinical covariates (sex, lesion laterality, seizure history, PSOM, time-since-stroke) were reserved for one-at-a-time sensitivity analyses (see Supplementary Materials). In addition to the primary model, exploratory analyses examined outcomes by language group (monolingual vs. simultaneous bilingual; both languages before age 3), following Leung et al.,^18^ to assess potential group-level trends. Analyses were conducted in R with permutation routines. We used Freedman–Lane permutation tests (B=10,000; covariate-adjusted) for both main effect of percentage of English exposure and for the omnibus interaction effects. We defined outcome domain families as Language (CELF subtests) and Cognition (Executive Function: D-KEFS; Nonverbal Reasoning: Matrix Reasoning). Within domain, we controlled multiplicity with Benjamini–Hochberg false discovery rate (FDR).^56^ Additional exploratory analyses with language groups (monolingual, bilingual) were conducted.

For interpretation, we report simple slopes of percentage of English within each category (β per +1 SD percentage of English exposure) and BH-adjusted pairwise differences of slopes. For omnibus tests, we report partial η² and its conversion to Cohen’s f². As descriptive benchmarks we use small/medium/large ≈ .01/.06/.14 for partial η² (≈ .02/.15/.35 for f²). For simple slopes and pairwise contrasts, effects are β per +1 SD in percentage English exposure, reported in each outcome’s transformed units (CELF = percentile rank; Wechsler Matrix Reasoning = scaled-score points) with 95% confidence intervals. As 1 SD of English exposure (z scored) in this sample corresponds to ∼26 percentage points, we also re-expressed slopes in terms of a +25-percentage point change in exposure to aid interpretability.

## Results

Between September 2024 and August 2025, 42 families were invited and 29 consented to the study. Of the 13 who did not participate, reasons included lack of interest (n=6), language barriers (n=2), or other logistical reasons (n=5). Our sample while modest is consistent with the size of prior pediatric stroke studies, where cohorts ranged from 20–40 participants.^57–59^

Mean age at stroke onset was 6.6 years (SD = 5.7), with stroke onset stratified across neonatal (≤28 days; n = 5), childhood (>28 days - 18 years; n = 13) and presumed perinatal (acquired perinatally but diagnosed after the neonatal period when neurological signs emerge; n = 11) groups. At assessment, participants were on average 13.4 years old (SD = 3.1) and 58.6% female. The sample was linguistically diverse including mono-, bi- and multilinguals, with similar English exposure across age at onset group: current English exposure averaged 69.2% (SD=26.4), one-third in each group reported ≥90% English exposure (functionally monolingual in daily contexts), and the average age of initial English exposure was between birth and early childhood. Full sample characteristics are presented in Table 2 (more details in Table S1).

**Table 2.**
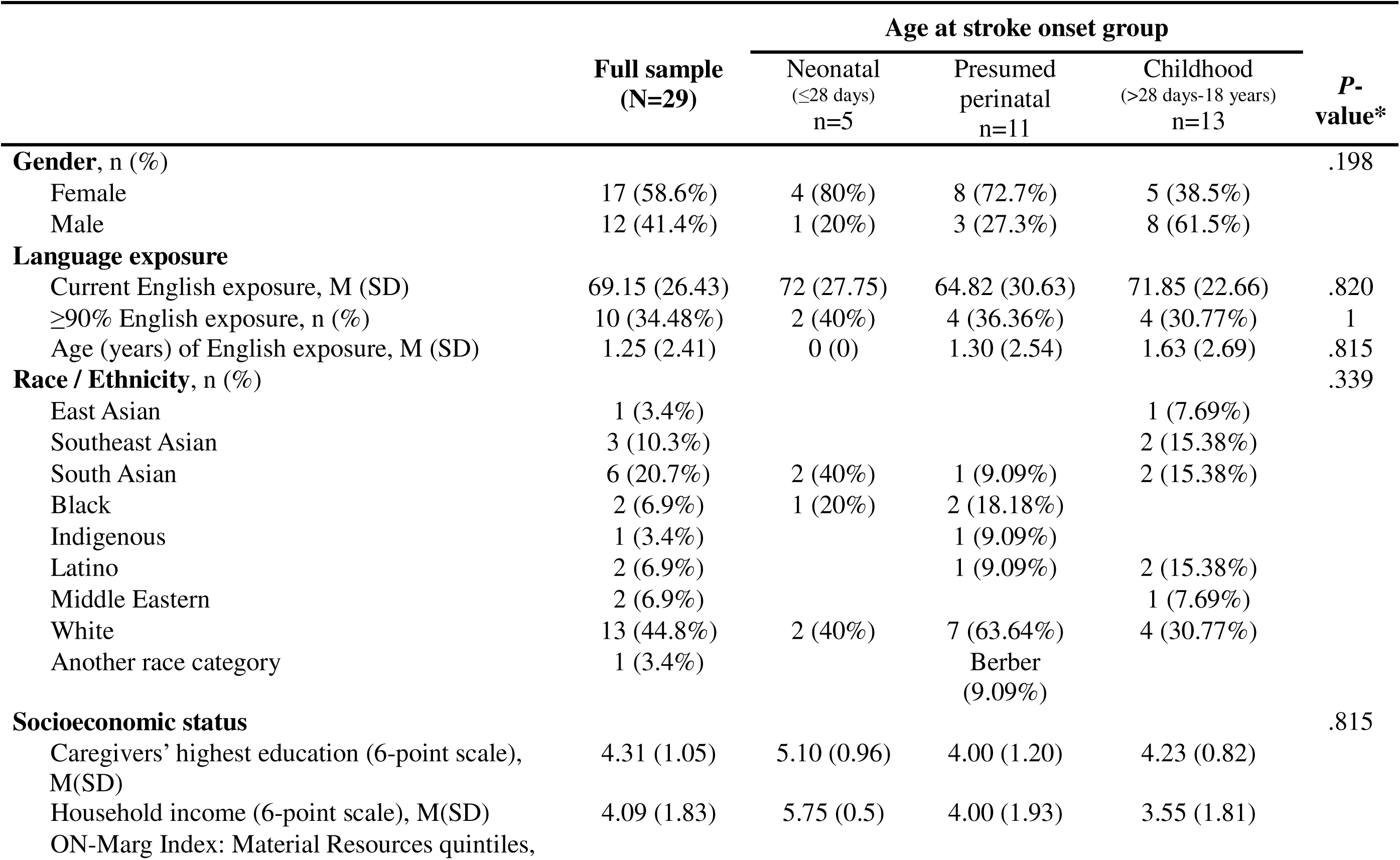
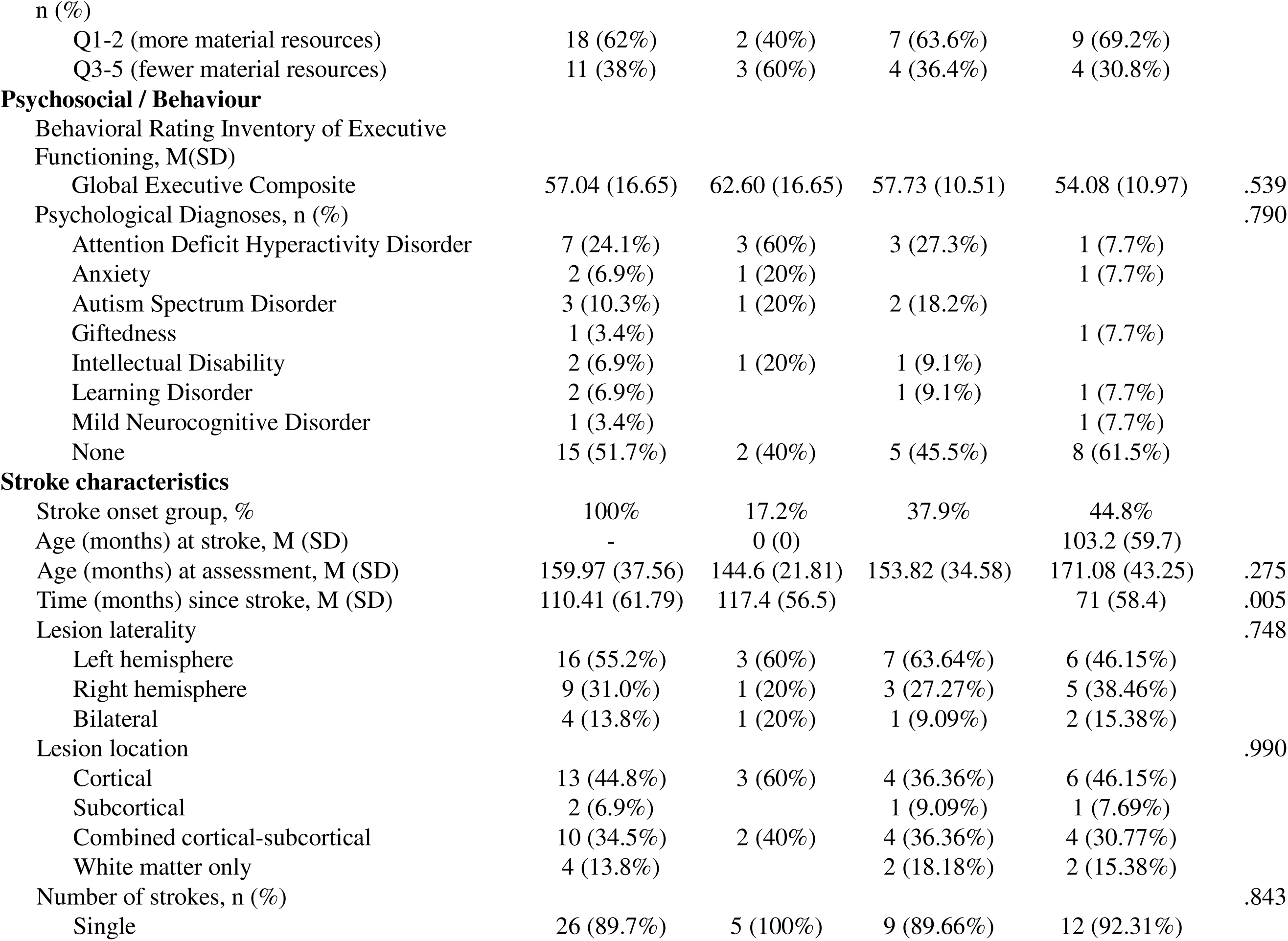
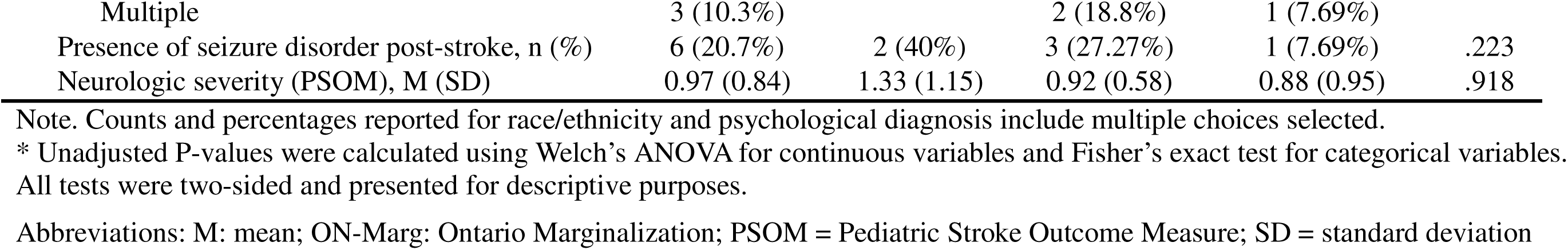
Clinical and demographic characteristics of the study participants.

|  | Full sample<br>(N=29) | Age at stroke onset group |  |  | P-value* |
| --- | --- | --- | --- | --- | --- |
|  |  | Neonatal<br>(≤28 days)<br>n=5 | Presumed<br>perinatal<br>n=11 | Childhood<br>(>28 days-18 years)<br>n=13 |  |
| **Gender, n (%)** |  |  |  |  | .198 |
| Female | 17 (58.6%) | 4 (80%) | 8 (72.7%) | 5 (38.5%) |  |
| Male | 12 (41.4%) | 1 (20%) | 3 (27.3%) | 8 (61.5%) |  |
| **Language exposure** |  |  |  |  |  |
| Current English exposure, M (SD) | 69.15 (26.43) | 72 (27.75) | 64.82 (30.63) | 71.85 (22.66) | .820 |
| ≥90% English exposure, n (%) | 10 (34.48%) | 2 (40%) | 4 (36.36%) | 4 (30.77%) | 1 |
| Age (years) of English exposure, M (SD) | 1.25 (2.41) | 0 (0) | 1.30 (2.54) | 1.63 (2.69) | .815 |
| **Race / Ethnicity, n (%)** |  |  |  |  | .339 |
| East Asian | 1 (3.4%) |  |  | 1 (7.69%) |  |
| Southeast Asian | 3 (10.3%) |  |  | 2 (15.38%) |  |
| South Asian | 6 (20.7%) | 2 (40%) | 1 (9.09%) | 2 (15.38%) |  |
| Black | 2 (6.9%) | 1 (20%) | 2 (18.18%) |  |  |
| Indigenous | 1 (3.4%) |  | 1 (9.09%) |  |  |
| Latino | 2 (6.9%) |  | 1 (9.09%) | 2 (15.38%) |  |
| Middle Eastern | 2 (6.9%) |  |  | 1 (7.69%) |  |
| White | 13 (44.8%) | 2 (40%) | 7 (63.64%) | 4 (30.77%) |  |
| Another race category | 1 (3.4%) |  | Berber<br>(9.09%) |  |  |
| **Socioeconomic status** |  |  |  |  | .815 |
| Caregivers' highest education (6-point scale),<br>M(SD) | 4.31 (1.05) | 5.10 (0.96) | 4.00 (1.20) | 4.23 (0.82) |  |
| Household income (6-point scale), M(SD) | 4.09 (1.83) | 5.75 (0.5) | 4.00 (1.93) | 3.55 (1.81) |  |
| ON-Marg Index: Material Resources quintiles, |  |  |  |  |  |
| n (%) |  |  |  |  |  |
| Q1-2 (more material resources) | 18 (62%) | 2 (40%) | 7 (63.6%) | 9 (69.2%) |  |
| Q3-5 (fewer material resources) | 11 (38%) | 3 (60%) | 4 (36.4%) | 4 (30.8%) |  |
| **Psychosocial / Behaviour** |  |  |  |  |  |
| Behavioral Rating Inventory of Executive Functioning, M(SD) |  |  |  |  |  |
| Global Executive Composite | 57.04 (16.65) | 62.60 (16.65) | 57.73 (10.51) | 54.08 (10.97) | .539 |
| Psychological Diagnoses, n (%) |  |  |  |  | .790 |
| Attention Deficit Hyperactivity Disorder | 7 (24.1%) | 3 (60%) | 3 (27.3%) | 1 (7.7%) |  |
| Anxiety | 2 (6.9%) | 1 (20%) |  | 1 (7.7%) |  |
| Autism Spectrum Disorder | 3 (10.3%) | 1 (20%) | 2 (18.2%) |  |  |
| Giftedness | 1 (3.4%) |  |  | 1 (7.7%) |  |
| Intellectual Disability | 2 (6.9%) | 1 (20%) | 1 (9.1%) |  |  |
| Learning Disorder | 2 (6.9%) |  | 1 (9.1%) | 1 (7.7%) |  |
| Mild Neurocognitive Disorder | 1 (3.4%) |  |  | 1 (7.7%) |  |
| None | 15 (51.7%) | 2 (40%) | 5 (45.5%) | 8 (61.5%) |  |
| **Stroke characteristics** |  |  |  |  |  |
| Stroke onset group, % | 100% | 17.2% | 37.9% | 44.8% |  |
| Age (months) at stroke, M (SD) | - | 0 (0) |  | 103.2 (59.7) |  |
| Age (months) at assessment, M (SD) | 159.97 (37.56) | 144.6 (21.81) | 153.82 (34.58) | 171.08 (43.25) | .275 |
| Time (months) since stroke, M (SD) | 110.41 (61.79) | 117.4 (56.5) |  | 71 (58.4) | .005 |
| Lesion laterality |  |  |  |  | .748 |
| Left hemisphere | 16 (55.2%) | 3 (60%) | 7 (63.64%) | 6 (46.15%) |  |
| Right hemisphere | 9 (31.0%) | 1 (20%) | 3 (27.27%) | 5 (38.46%) |  |
| Bilateral | 4 (13.8%) | 1 (20%) | 1 (9.09%) | 2 (15.38%) |  |
| Lesion location |  |  |  |  | .990 |
| Cortical | 13 (44.8%) | 3 (60%) | 4 (36.36%) | 6 (46.15%) |  |
| Subcortical | 2 (6.9%) |  | 1 (9.09%) | 1 (7.69%) |  |
| Combined cortical-subcortical | 10 (34.5%) | 2 (40%) | 4 (36.36%) | 4 (30.77%) |  |
| White matter only | 4 (13.8%) |  | 2 (18.18%) | 2 (15.38%) |  |
| Number of strokes, n (%) |  |  |  |  | .843 |
| Single | 26 (89.7%) | 5 (100%) | 9 (89.66%) | 12 (92.31%) |  |
| Multiple | 3 (10.3%) |  | 2 (18.8%) | 1 (7.69%) |  |
| Presence of seizure disorder post-stroke, n (%) | 6 (20.7%) | 2 (40%) | 3 (27.27%) | 1 (7.69%) | .223 |
| Neurologic severity (PSOM), M (SD) | 0.97 (0.84) | 1.33 (1.15) | 0.92 (0.58) | 0.88 (0.95) | .918 |

Note. Counts and percentages reported for race/ethnicity and psychological diagnosis include multiple choices selected.
\* Unadjusted P-values were calculated using Welch's ANOVA for continuous variables and Fisher's exact test for categorical variables. All tests were two-sided and presented for descriptive purposes.
Abbreviations: M: mean; ON-Marg: Ontario Marginalization; PSOM = Pediatric Stroke Outcome Measure; SD = standard deviation

Outcome data was largely complete (see Table S2 for test availability). Missing data primarily reflected administrative exclusions (e.g., time constraints, omitted for other reasons) or ineligibility (e.g., age-related criteria for the CELF-5 M). For measures deemed too difficult for certain participants, the worst-rank procedure was applied, and scores were converted to sample-relative percentiles (0-1; higher=better).

### Descriptive results

As shown in Table 3, language and cognitive outcomes summarized as within-sample percentile rank (0.00–1.00, higher = better; median [IQR], *n* available) were broadly similar across age at stroke groups for CELF-5 Formulated Sentences and Making Inferences (medians ≈0.45–0.46). On Multiple Meanings, the neonatal group showed a higher median (59^th^ percentile) than other groups (childhood, 36^th^ percentile; presumed, 41^st^ percentile). In cognitive tasks, the presumed onset group tended to be higher on Inhibition (58^th^ percentile) and Inhibition/Switching (72^nd^ percentile) than childhood (30^th^ and 42^nd^ percentile respectively), whereas childhood was higher on Number–Letter Switching (51^st^ vs. presumed 31^st^). For Wechsler Matrix Reasoning scaled scores, the stroke sample performed slightly below the norm; the neonatal group had a lower median than childhood and presumed (median 6 vs. both 9).

**Table 3.** Performance on language and cognitive outcomes across age at stroke onset groups (median [IQR]; n)

| Domain | Outcome | Full sample | Age at stroke onset group |  |  |
| --- | --- | --- | --- | --- | --- |
|  |  |  | Neonatal<br>(≤28 days) | Presumed<br>Perinatal | Childhood<br>(29 days-18 years) |
| Language | CELF-5<br>Formulated Sentences<br>(percentile rank) | 0.46 [0.15–0.69];<br>n=29 | 0.46 [0.15–0.90];<br>n=5 | 0.46 [0.15–0.57];<br>n=11 | 0.46 [0.15–0.62];<br>n=13 |
|  | CELF-5 M<br>Making Inferences<br>(percentile rank) | 0.45 [0.12–0.69];<br>n=25 | 0.45 [0.19–0.69];<br>n=5 | 0.45 [0.02–0.60];<br>n=9 | 0.45 [0.15–0.75];<br>n=11 |
|  | CELF-5 M<br>Multiple Meanings<br>(percentile rank) | 0.41 [0.23–0.68];<br>n=26 | 0.59 [0.23–0.80];<br>n=5 | 0.41 [0.02–0.59];<br>n=9 | 0.36 [0.23–0.71];<br>n=12 |
| Cognition | D-KEFS Inhibition<br>(percentile rank) | 0.46 [0.16–0.70];<br>n=28 | 0.46 [0.24–0.46];<br>n=5 | 0.58 [0.46–0.70];<br>n=10 | 0.30 [0.12–0.70];<br>n=13 |
|  | D-KEFS<br>Inhibition/Switching<br>(percentile rank) | 0.42 [0.14–0.72];<br>n=28 | 0.58 [0.14–0.58];<br>n=5 | 0.72 [0.28–0.82];<br>n=10 | 0.42 [0.14–0.58];<br>n=13 |
|  | D-KEFS<br>Number–Letter Switching<br>(percentile rank) | 0.39 [0.14–0.76];<br>n=29 | 0.39 [0.24–0.51];<br>n=5 | 0.31 [0.19–0.72];<br>n=11 | 0.51 [0.14–0.76];<br>n=13 |
|  | Wechsler Matrix Reasoning<br>(scaled score) | 9.00 [6.00–10.00];<br>n=29 | 6.00 [4.00–9.00];<br>n=5 | 9.00 [7.50–11.00];<br>n=11 | 9.00 [7.00–10.00];<br>n=13 |

*Note.* Percentile rank outcomes are within-sample, not population-based norms (0–1, higher = better). Matrix Reasoning is reported as scaled scores (norm mean = 10, SD = 3).
Abbreviations: CELF-5 = Clinical Evaluation of Language Fundamentals–Fifth Edition; D-KEFS = Delis–Kaplan Executive Function System; IQR = interquartile range.

### Permutation testing results

Analyses operationalized linguistic diversity as percentage of English exposure (z-scored); results are interpreted in terms of this framing (i.e., greater linguistic diversity = lower percentage of English exposure).

#### Main effect of English exposure

Across outcomes, there were no domain-wise main effects of linguistic diversity after BH correction (Table S3-4; Language p_perm_ ≥ .200; FDR q ≥ .475; Cognition p_perm_ ≥ .409; FDR q ≥ .785).

#### Interaction model (percentage of English exposure × age at stroke)

##### Language

By contrast, linguistic diversity did interact with age at stroke for three CELF outcomes (Formulated Sentences: p_perm_=.004, q=.012, partial η²=.39; Multiple Meanings: p_perm_ =.008, q=.012, partial η²=.39; Making Inferences: p_perm_ =.053, q=.053, partial η²=.28; Table 4).

**Table 4.** Omnibus interaction tests for language outcomes (model: Outcome ∼ linguistic diversity (z) × age at stroke + SES composite (z); permutation p from Freedman–Lane; BH–FDR within the language domain).

| Outcome | N | F-value | df | $p_{\text{perm}}$ | $q$<br>(BH) | partial $\eta^2$ | $f^2$ |
| --- | --- | --- | --- | --- | --- | --- | --- |
| CELF-5: Formulated Sentences<br>(percentile rank) | 29 | 6.95 | 2, 22 | **.004** | **.012** | 0.387 | 0.632 |
| CELF-5 M: Multiple Meanings<br>(percentile rank) | 26 | 6.08 | 2, 19 | **.008** | **.012** | 0.390 | 0.640 |
| CELF-5 M: Making Inferences<br>(percentile rank) | 25 | 3.44 | 2, 18 | <u>.053</u> | <u>.053</u> | 0.277 | 0.382 |

*Note.* Significant results are bolded. partial $\eta^2$ benchmarks: small .01; medium .06; large .14. Abbreviations: CELF-5 (M): Clinical Evaluation of Language Fundamentals, Fifth Edition (Metalinguistics); df: degrees of freedom; BH: Benjamini–Hochberg procedure.

Simple slopes and pairwise testing (Figure 1, Full slope estimates in S5) showed that in the neonatal onset group, greater linguistic diversity was significantly associated with higher scores on Formulated Sentences (β = −0.317, 95% CI [−0.612, −0.023]), equivalent to ∼0.30 SD higher scores per 25-percentage-points greater linguistic diversity. Trends were also seen on the CELF Metalinguistics subtests. In the childhood group, associations were near-zero. By contrast, in the presumed stroke group, greater linguistic diversity was significantly linked to lower scores across subtests, with steeper slopes than in the neonatal and childhood onset groups for Formulated Sentences and Multiple Meanings.

**Figure 1.**
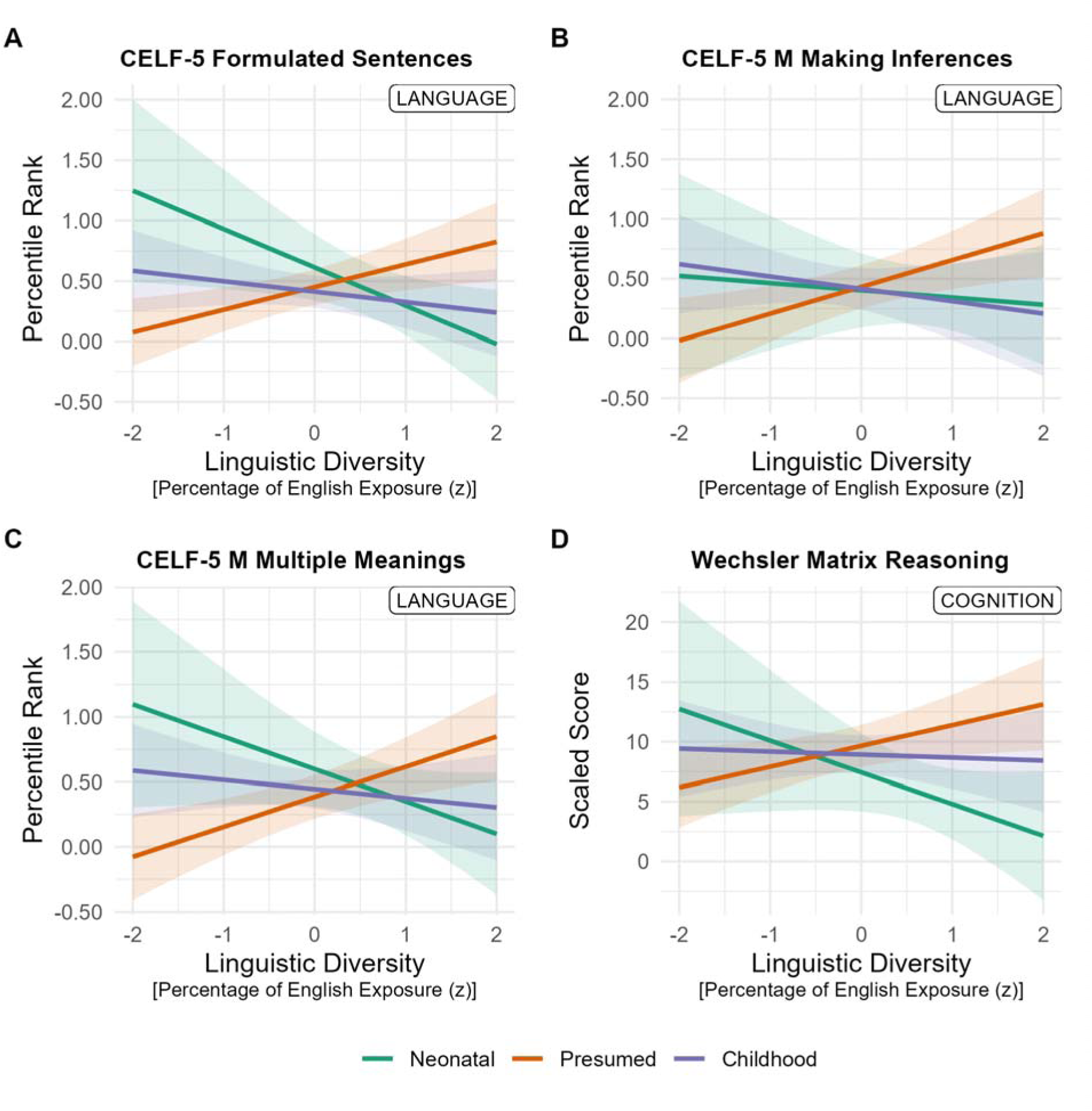
Predicted performance based on linguistic diversity by age at stroke (Neonatal, Presumed, Childhood interactions. Higher percentage of English exposure (z-scored) indicates lower linguistic diversity in the environment. Panels A–C (Language) show CELF outcomes— Formulated Sentences, Making Inferences, Multiple Meanings—on sample-relative percentile ranks (higher = better); Panel D (Cognition) shows Wechsler Matrix Reasoning (scaled score). Lines are model-based predictions from linear models with a percentage of English exposure (z-scored) × age-at-stroke interaction (SES z covariate); shaded bands indicate 95% CIs.

##### Cognition (Executive function, Nonverbal Reasoning)

For Matrix Reasoning, the age × exposure omnibus effect patterned significantly (p_perm_=.056, q=.226, partial η²=.23) with the same directionality: positive in neonatal, ∼0 in childhood, negative in presumed. D-KEFS outcomes showed no reliable interactions (q≥.37; Table 5.).

**Table 5.**
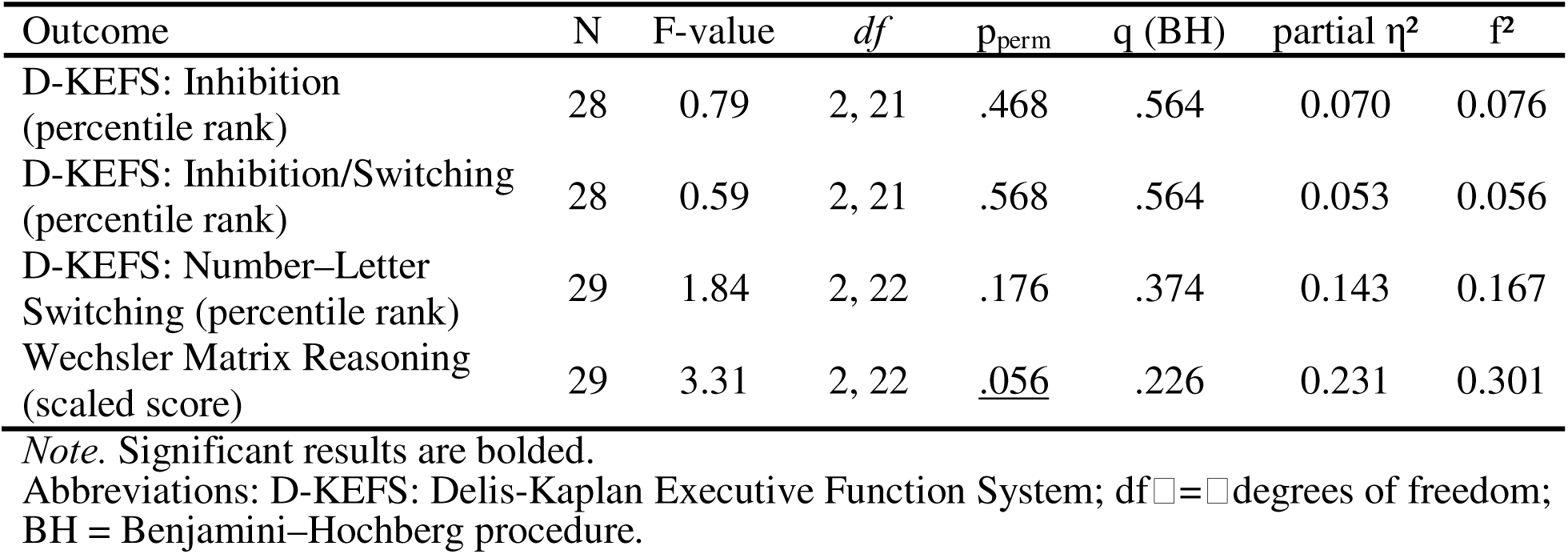
Freedman–Lane permutation test model examining impact of linguistic diversity, age at stroke and its interaction on cognitive outcomes, controlling for SES.

Simple slopes and pairwise testing (**Error! Reference source not found.**, Table S6) showed that in the neonatal onset group, greater linguistic diversity was associated with better Matrix Reasoning scores (β = −2.65, 95% CI [−6.16, 0.86]; ≈ 2.55 points increase per 25 percentage points greater linguistic diversity), although the confidence interval includes 0, underscoring imprecision of the finding. In the childhood group, the association neared zero (β = −0.25, 95% CI [−2.28, 1.78]). By contrast, among children who were of the presumed onset group, greater linguistic diversity was associated with worse scores (β = 1.73, 95% CI [0.07, 3.40]; ≈1.66 points decrease); the slope for this association was steeper than neonatal (Δβ = 4.38, q = .080) and childhood groups (Δβ = 1.98, q = .195), though neither contrast survived BH correction.

### Sensitivity and exploratory analyses

To assess the robustness of our primary models, we conducted one-at-a-time sensitivity analyses adding clinical covariates (sex, lesion laterality, post-stroke seizure, PSOM, and time since stroke). Results were largely consistent with the primary findings: no new significant main effects of English exposure emerged after adjustment; the linguistic diversity × age at stroke interactions remained robust to most covariates (see Supplementary Results and Tables S7-8 for full details).

Exploratory analyses by categorical language group (monolingual vs. simultaneous bilingual) yielded patterns largely consistent with the continuous linguistic diversity model (see Supplementary Materials). For CELF Formulated Sentences, bilinguals outperformed monolinguals in the neonatal group (Δ=0.56, *p*=.033, Table S12), with a comparable trend in the childhood group (*p*=.066, Table S12). In contrast, on Matrix Reasoning, monolinguals performed better within the presumed-onset group (*p*=.005, Table S12). Other measures (e.g., CELF Making Inferences, D-KEFS) showed no additional group differences. Given the small sample size, these categorical findings are best viewed as exploratory and complementary to the continuous analyses.

## Discussion

This study examined how linguistic diversity and stroke-related factors relate to language and cognitive outcomes in a linguistically diverse ischemic stroke cohort, addressing key drivers of health inequities in neurologic care. Our analyses modelled linguistic diversity (percentage of English exposure), controlling for SES. There were no main effects of linguistic diversity across domains. However, associations varied by age at stroke onset. Children with childhood-onset stroke (onset between 29 days and 18 years) showed near-zero associations, suggesting that this group may be relatively stable across differences in linguistic environment. In contrast, a task-specific (strongest in expressive language) advantage emerged in the neonatal group (onset ≤28 days), where greater linguistic diversity related to better language scores. For cognition, Matrix Reasoning showed a similar trend to the language finding, though did not survive multiple testing correction. By contrast, the presumed perinatal group showed an opposite pattern: children with greater linguistic diversity performed worse on language outcomes. We observed no group differences in executive functioning.

These findings extend prior retrospective longitudinal work in pediatric stroke in bilinguals^18^ and align with evidence from other developmental disorders, where bilingual children often perform as well as or better than monolingual peers.^41,42^ At the same time, the absence of consistent advantages across all domains - including no reliable effects on executive functioning in this cohort - and stroke types reflects the heterogeneity of bilingualism and their ongoing debated existence in the bilingualism literature.^36^ The patients’ task profile in this study is inconsistent with evidence that bilingual effects are more likely found on control-intensive measures, such as executive functioning. The findings reinforce that age at injury, the amount and type of language exposure, and sociolinguistic context jointly influence developmental outcomes, underscoring the need to move beyond categorical bilingual-monolingual distinctions.^43,60^

Although neonatal and presumed perinatal strokes are close in developmental timing, their bilingualism–outcome associations diverged. One possibility to consider is the different way the strokes are ascertained and their respective phenotypes. Neonatal strokes are typically diagnosed at the time of injury because of acute symptoms,^61^ whereas presumed perinatal strokes are diagnosed later, when chronic sequelae (e.g., hemiparesis, early hand preference, global delay) prompt neuroimaging. As a result, this group may over-represent children with persistent, functionally significant injuries.^57,62–64^ In other studies that included neonatal and presumed perinatal patients, Ghotra and colleagues also saw presumed perinatal stroke participants with the highest morbidity including lowest global and motor abilities and lowest independence in daily activities.^63^ Additionally, lesion characteristics can influence outcomes independently of bilingualism and bias comparisons if not accounted for.^22,29^ In our sample, lesion descriptors (e.g., location and laterality) did not differ systematically between neonatal and presumed groups, and sensitivity analyses adjusting for seizure history yielded similar patterns, suggesting the divergence is not explained by these factors. Nevertheless, retrospective identification of presumed cases may still introduce group-definition bias, which likely captures a distinct neurodevelopmental and clinical profile.^64^ Moreover, presumed cases are often excluded in prior pediatric stroke studies,^15,17,18^ limiting predictive frameworks for this group in interactions with bilingual exposure. The distinct behavioral profile observed here highlights the importance of including presumed cases in future research, ideally with richer lesion and severity characterization. Given the exploratory nature of analyses in this subgroup, the presumed perinatal findings should not be interpreted as evidence that bilingual exposure causally harms outcomes, but rather as highlighting the need for replication with comprehensive clinical characterization.

From a theoretical perspective, these findings underscore the role of developmental timing and neuroplasticity. Early injury, particularly in the perinatal period (e.g., presumed, neonatal), may amplify the influence of the language environment while neural systems are still consolidating, whereas later childhood stroke occurs after more robust networks are established, allowing these systems to buffer against differences in input and thus reducing the influence of language exposure.^22,65^ This pattern aligns with neuroplasticity models, which emphasize that the timing of injury shapes the extent and nature of functional reorganization.^66^ Importantly, in the neonatal and childhood-onset group, which together comprised the majority of the sample, there was no evidence that bilingual exposure negatively affected outcomes. The presumed perinatal findings remain inconclusive given the diagnostic and clinical complexities of this subgroup. In some contexts, such as neonatal stroke, diverse language exposure may facilitate adaptive reorganization supporting language, whereas in others, outcomes may depend more strongly on lesion characteristics and developmental timing.

These findings must also be interpreted within the broader environmental context in which linguistic diversity operates. As noted earlier, SES predicts post-stroke cognitive and language outcomes^26,28^ and was included as a covariate in our models to isolate linguistic diversity effects. This approach is supported by meta-analytic evidence that many reported bilingual advantages in executive function disappear when SES is objectively measured and controlled.^67^ The relationship between bilingualism and SES appears to vary by population and domain: neurotypical studies have found independent, additive effects,^68,69^ while in some clinical and neurotypical populations, bilingualism may confer greater advantages for children from lower SES.^70,71^ Our design prioritized age at stroke as the primary moderator while controlling for SES, but future studies with larger pediatric stroke samples should explicitly test whether linguistic diversity confers differential benefits across socioeconomic contexts, with the goal of informing equity-oriented interventions that account for both language environment and socioeconomic factors in supporting children post-stroke.

### Clinical implications

These findings challenge outdated assumptions that bilingual exposure uniformly pose a burden. For children with neonatal and childhood-onset stroke, these results underscore the need for culturally and linguistically responsive practices that affirm, rather than restrict, children’s linguistic environments.^72–74^ The presumed perinatal group showed a different pattern, but given the diagnostic ambiguity, clinical heterogeneity, and exploratory nature of these analyses, this finding should not be taken as evidence to restrict bilingual exposure without further replication and mechanistic investigation. Clinical assessments should account for linguistic diversity, as applying monolingual norms to bilingual children can lead to misdiagnosis and underestimation of abilities. Close monitoring of language development while supporting home/heritage languages is important across stroke-timing profiles, with individualized attention to dominant language needs in early strokes and functional cross-linguistic communication in later-onset cases.

### Limitations

Several limitations should be considered. The sample size was modest, with uneven subgroup distributions, reducing statistical power, and limiting precision of slope estimates. Permutation-based inference can be used as a model for increased confidence in the robustness of the observed interactions amongst smaller samples, but replication in larger cohorts will be essential. The cross-sectional design also limits inference about trajectories following this timepoint and potential confounders such as comorbidities, other neurological factors (e.g., lesion size/volume, lateralization of language), rehabilitation intensity and literacy practices were not fully accounted for.

Recruitment-related selection bias may have led to underrepresentation of families facing language barriers or logistical and socioeconomic constraints when navigating English-dominant research settings. Future research could employ community-engaged recruitment and offer multilingual consent and assessment to reduce participation barriers. Generalizability is constrained by single-site design and sociocultural context, as language exposure and support for bilingualism varies across regions. Larger, multi-site samples would enable multivariate testing of environmental factors simultaneously.

Measurement limitations include that percentage of English exposure captures quantity but not necessarily the proficiency, balance, code-switching, or context of use. For pragmatic reasons, we were unable to directly assess non-English abilities with a standardized test battery. Higher English exposure may advantage performance on English-administered language tasks. Cognitive outcomes were derived from a mix of standardized and sample-relative measures, and parent-reported executive functioning may reflect caregiver perceptions as much as child performance.

Finally, the presumed perinatal group represents a diagnostically and clinically distinct population, identified retrospectively and potentially overrepresenting more severe or atypical injuries. The negative association between linguistic diversity and outcomes in this group may reflect unmeasured confounders (e.g., injury severity, pre-stroke development, access to early intervention) or reverse causation and should be considered exploratory pending replication in prospectively characterized cohorts.

## Conclusion

In conclusion, for children with neonatal and childhood-onset stroke, bilingual environments do not impede and may support language and cognitive development. Results for the presumed perinatal group were more complex and require replication with richer clinical characterization before drawing definitive clinical conclusions. Overall, these findings challenge the misconception that bilingual exposure uniformly compromises development post-injury and advocate for clinical approaches that affirm linguistic diversity while attending to individual developmental and neurological contexts. Future work should incorporate direct measures of proficiency and naturalistic language use to provide a more nuanced understanding of bilingual exposure. Translation of these findings into clinical practice guidelines is a crucial next step, ensuring that rehabilitation providers are equipped to support all children with evidence-based, culturally responsive approaches.

## Acknowledgements

We thank the children and families who participated in this study. We are grateful to the clinical team at The Hospital for Sick Children for their support, in particular Ms. Elena Sanjuan and Ms. Ana Radmilovic. We also acknowledge Ms. Victoria Gotcheva for her help in optimizing data collection workflows and Dr. Tijana Simic for their help in reviewing an earlier version of this manuscript and.

## Author Contributions

KIL: Conceptualization, Methodology, Formal Analysis, Investigation, Data Curation, Formal analysis, Visualization, Writing – Original Draft, Writing – Review & Editing, Funding acquisition

RW: Conceptualization, Methodology, Investigation, Resources, Writing – Review & Editing, Supervision

ND: Conceptualization, Methodology, Resources, Writing – Review & Editing, Supervision

ER: Conceptualization, Methodology, Writing – Review & Editing, Supervision

MM: Conceptualization, Methodology, Resources, Writing – Review & Editing, Supervision, Funding Acquisition

## Statements and Declarations

### Ethical Considerations

Ethical approval for this study was obtained from The Hospital for Sick Children Research Ethics Board (REB# 1000081176).

### Consent to Participate

Written informed consent was obtained from all caregivers and assent was obtained from youth participants prior to participation in this study.

### Consent for Publication

Not applicable

### Data Availability

The data that support the findings of this study are available in the Supplementary Materials.

## References

1. Champigny CM, Feldman SJ, Westmacott R, et al. Adjusting to life after pediatric stroke: A qualitative study. Dev Med Child Neurol. 2023;65(10):1357–1365. doi:10.1111/dmcn.15556

2. Gao L, Lim M, Nguyen D, et al. The incidence of pediatric ischemic stroke: A systematic review and meta-analysis. Int J Stroke. 2023;18(7):765–772. doi:10.1177/17474930231155336

3. Rohner A, Gutbrod K, Kohler B, et al. Health-Related Quality of Life in Young Adults Following Pediatric Arterial Ischemic Stroke. Stroke. 2020;51(3):952–957. doi:10.1161/STROKEAHA.119.027622

4. Kim CT, Han J, Kim H. Pediatric stroke recovery: a descriptive analysis. Arch Phys Med Rehabil. 2009;90(4):657–662. doi:10.1016/j.apmr.2008.10.016

5. Kirton A, Westmacott R, Deveber G. Pediatric stroke: Rehabilitation of focal injury in the developing brain. NeuroRehabilitation. 2007;22(5):371–382. doi:10.3233/NRE-2007-22504

6. Westmacott R, Askalan R, Macgregor D, Anderson P, Deveber G. Cognitive outcome following unilateral arterial ischaemic stroke in childhood: effects of age at stroke and lesion location. Dev Med Child Neurol. 2010;52(4):386–393. doi:10.1111/j.1469-8749.2009.03403.x

7. Westmacott R, McDonald KP, Roberts SD, et al. Predictors of cognitive and academic outcome following childhood subcortical stroke. Dev Neuropsychol. 2018;43(8):708–728. doi:10.1080/87565641.2018.1522538

8. Gilardone G, Viganò M, Cassinelli D, et al. Post-stroke acquired childhood aphasia. A scoping review. Child Neuropsychol. 2023;29(8):1268–1293. doi:10.1080/09297049.2022.2156992

9. Greenham M, Anderson V, Mackay MT. Improving cognitive outcomes for pediatric stroke. Curr Opin Neurol. 2017;30(2):127–132. doi:10.1097/WCO.0000000000000422

10. O’Keeffe F, Murphy O, Ganesan V, King J, Murphy T. Neuropsychological outcome following childhood stroke – a review. Brain Inj. 2017;31(12):1575–1589. doi:10.1080/02699052.2017.1332782

11. Williams TS, McDonald KP, Roberts SD, Dlamini N, deVeber G, Westmacott R. Prevalence and predictors of learning and psychological diagnoses following pediatric arterial ischemic stroke. Dev Neuropsychol. 2017;42(5):309–322. doi:10.1080/87565641.2017.1353093

12. Ferriero DM, Fullerton HJ, Bernard TJ, et al. Management of Stroke in Neonates and Children: A Scientific Statement From the American Heart Association/American Stroke Association. Stroke. 2019;50(3). doi:10.1161/STR.0000000000000183

13. Roach ES, Golomb MR, Adams R, et al. Management of Stroke in Infants and Children: A Scientific Statement From a Special Writing Group of the American Heart Association Stroke Council and the Council on Cardiovascular Disease in the Young. Stroke. 2008;39(9):2644–2691. doi:10.1161/STROKEAHA.108.189696

14. Abgottspon S, Thaqi Q, Steiner L, et al. Effect of Age at Pediatric Stroke on Long-term Cognitive Outcome. Neurology. 2022;98(7). doi:10.1212/WNL.0000000000013207

15. Felling RJ, Rafay MF, Bernard TJ, et al. Predicting recovery and outcome after pediatric stroke: results from the International Pediatric Stroke Study. Ann Neurol. 2020;87(6):840–852. doi:10.1002/ana.25718

16. Golomb MR, MacGregor DL, Domi T, et al. Presumed pre or perinatal arterial ischemic stroke: risk factors and outcomes. Ann Neurol. 2001;50(2):163–168. doi:10.1002/ana.1078

17. Sullivan AW, Johnson MK, Boes AD, Tranel D. Implications of age at lesion onset for neuropsychological outcomes: A systematic review focusing on focal brain lesions. Cortex. 2023;163:92–122. doi:10.1016/j.cortex.2023.03.002

18. Leung KI, Dlamini N, Westmacott R, Molnar M. Language and Cognitive Outcomes Following Ischemic Stroke in Children With Monolingual and Bilingual Exposure. J Child Neurol. 2023;38(6-7). doi:10.1177/08830738231171466

19. Fuentes A, Deotto A, Desrocher M, deVeber G, Westmacott R. Determinants of cognitive outcomes of perinatal and childhood stroke: A review. Child Neuropsychol. 2016;22(1):1–38. doi:10.1080/09297049.2014.969694

20. Krivitzky LS, Westmacott R, Boada R, Sepeta L, Reppert L, Mrakotsky C. Recent Advances in Neuropsychological Outcomes and Intervention in Pediatric Stroke. Stroke. 2022;53(12):3780–3789. doi:10.1161/STROKEAHA.122.037294

21. Pabst L, Hoyt CR, Felling RJ, et al. Neuroimaging and Neurological Outcomes in Perinatal Arterial Ischemic Stroke: A Systematic Review and Meta-Analysis. Pediatr Neurol. 2024;157:19–28. doi:10.1016/j.pediatrneurol.2024.04.029

22. Anderson V, Darling S, Mackay M, et al. Cognitive resilience following paediatric stroke: Biological and environmental predictors. Eur J Paediatr Neurol. 2020;25:52–58. doi:10.1016/J.EJPN.2019.11.011

23. Faigle R, Towfighi A. Advances in the understanding of social determinants of health in stroke. Stroke. 2024;55(6):1680–1682. doi:10.1161/STROKEAHA.124.041733

24. Greenham M, Hearps S, Gomes A, et al. Environmental contributions to social and mental health outcomes following pediatric stroke. Dev Neuropsychol. 2015;40(6):348–362. doi:10.1080/87565641.2015.1095191

25. Gordon AL. Functioning and disability after stroke in children: using the ICF-CY to classify health outcome and inform future clinical research priorities. Dev Med Child Neurol. 2014;56(5):434–444. doi:10.1111/dmcn.12336

26. Jordan LC, Hills NK, Fox CK, et al. Socioeconomic determinants of outcome after childhood arterial ischemic stroke. Neurology. 2018;91(6):e509–e516. doi:10.1212/WNL.0000000000005946

27. Pai AM, To T, deVeber GA, et al. Health Inequity and Time From Pediatric Stroke Onset to Arrival. Stroke. 2024;55(5):1299–1307. doi:10.1161/STROKEAHA.123.045411

28. Bartha-Doering L, Gleiss A, Knaus S, Schmook MT, Seidl R. Influence of socioeconomic status on cognitive outcome after childhood arterial ischemic stroke. Dev Med Child Neurol. 2021;63(4):465–471. doi:10.1111/dmcn.14779

29. Champigny CM, Feldman SJ, Beribisky N, et al. Predictors of neurocognitive outcome in pediatric ischemic and hemorrhagic stroke. Child Neuropsychol. 2024;30(3):444–461. doi:10.1080/09297049.2023.2213461

30. Grosjean F. Bilingualism: A short introduction. Psycholinguist Biling. 2013;2(5):5–25.

31. Hakuta K. Mirror of Language. The Debate on Bilingualism. ERIC; 1986.

32. Peal E, Lambert WE. The relation of bilingualism to intelligence. Psychol Monogr Gen Appl. 1962;76(27):1. doi:10.1037/h0093840

33. Kay Raining-Bird E, Lamond E, Holden J. Survey of bilingualism in autism spectrum disorders. Int J Lang Commun Disord. 2012;47(1):52–64. doi:10.1111/J.1460-6984.2011.00071.X

34. Marinova-Todd SH, Colozzo P, Mirenda P, et al. Professional practices and opinions about services available to bilingual children with developmental disabilities: An international study. J Commun Disord. 2016;63:47–62. doi:10.1016/J.JCOMDIS.2016.05.004

35. Bialystok E. Beyond executive functioning: rethinking the impact of bilingualism. Trends Cogn Sci. 2025;29(3):220–221. doi:10.1016/j.tics.2024.12.005

36. Paap K. The Bilingual Advantage Debate. Handb Neurosci Multiling. Published online February 19, 2019:701–735. doi:10.1002/9781119387725.CH34

37. Bialystok E, Craik FIM. Cognitive and linguistic processing in the bilingual mind. Curr Dir Psychol Sci. 2010;19(1). doi:10.1177/0963721409358571

38. Green DW, Abutalebi J. The adaptive control hypothesis. J Cogn Psychol. 2013;25(5):515–530. doi:10.1080/20445911.2013.796377

39. Bialystok E. Bilingualism modifies cognition through adaptation, not transfer. Trends Cogn Sci. 2024;28(11):987–997. doi:10.1016/j.tics.2024.07.012

40. Leung KI, Molnar M. Examining Neurodiversity in Bilingual Development Research: Recent Insights Through an Equity, Diversity, and Inclusion Lens. Int J Lang Commun Disord. 2025;60(5):e70100. doi:10.1111/1460-6984.70100

41. Kay-Raining Bird E, Genesee F, Verhoeven L. Bilingualism in children with developmental disorders: A narrative review. J Commun Disord. 2016;63:1–14. doi:10.1016/j.jcomdis.2016.07.003

42. Uljarević M, Katsos N, Hudry K, Gibson JL. Practitioner Review: Multilingualism and neurodevelopmental disorders–an overview of recent research and discussion of clinical implications. J Child Psychol Psychiatry. 2016;57(11):1205–1217. doi:10.1111/jcpp.12596

43. Kremin LV, Byers-Heinlein K. Why not both? Rethinking categorical and continuous approaches to bilingualism. Int J Biling. 2021;25(6):1560–1575. doi:10.1177/13670069211031986

44. Feldman SJ, Beslow LA, Felling RJ, et al. Consensus-Based Evaluation of Outcome Measures in Pediatric Stroke Care: A Toolkit. Pediatr Neurol. Published online January 19, 2023. doi:10.1016/J.PEDIATRNEUROL.2023.01.009

45. Wiig EH, Secord WA, Semel E. Clinical Evaluation of Language Fundamentals. Fifth Edition. Pearson; 2013.

46. Wiig EH, Secord WA. Clinical Evaluation of Language Fundamentals Metalinguistics. Fifth Edition. Pearson; 2014.

47. Wechsler D. Wechsler Intelligence Scale for Children. Fifth Edition. Pearson; 2014.

48. Wechsler D. Wechsler Adult Intelligence Scale. Fourth Edition. Pearson; 2008.

49. Delis DC, Kaplan E, Kramer J. Delis-Kaplan Executive Function System. Psychological Corporation; 2001.

50. Yu AY, Smith EE, Krahn M, et al. Association of neighborhood-level material deprivation with health care costs and outcome after stroke. Neurology. 2021;97(15):e1503–e1511. doi:10.1212/WNL.0000000000012676

51. Marian V, Blumenfeld HK, Kaushanskaya M. The Language Experience and Proficiency Questionnaire (LEAP-Q): Assessing Language Profiles in Bilinguals and Multilinguals. J Speech Lang Hear Res. 2007;50(4):940–967. doi:10.1044/1092-4388(2007/067)

52. Gioia G, Isquith PK, Guy S, Kenworthy L. Behavior Rating Inventory of Executive Function. Second Edition. Psychological Assessment Resources, Inc; 2015.

53. Lang JM, Connell CM. Development and validation of a brief trauma screening measure for children: The Child Trauma Screen. Psychol Trauma Theory Res Pract Policy. 2017;9(3):390–398. doi:10.1037/tra0000235

54. Murphy JM, Bergmann P, Chiang C, et al. The PSC-17: Subscale Scores, Reliability, and Factor Structure in a New National Sample. Pediatrics. 2016;138(3):e20160038. doi:10.1542/peds.2016-0038

55. Kitchen L, Westmacott R, Friefeld S, et al. The pediatric stroke outcome measure: a validation and reliability study. Stroke. 2012;43(6):1602–1608. doi:10.1161/STROKEAHA.111.639583

56. Benjamini Y, Hochberg Y. Controlling the False Discovery Rate: A Practical and Powerful Approach to Multiple Testing. J R Stat Soc Ser B Stat Methodol. 1995;57(1):289–300. doi:10.1111/j.2517-6161.1995.tb02031.x

57. Bosenbark DD, Krivitzky L, Ichord R, Jastrzab L, Billinghurst L. Attention and executive functioning profiles in children following perinatal arterial ischemic stroke. Child Neuropsychol. 2018;24(1):106–123. doi:10.1080/09297049.2016.1225708

58. Heimgärtner M, Gschaidmeier A, Schnaufer L, Staudt M, Wilke M, Lidzba K. The long-term negative impact of childhood stroke on language. Front Pediatr. 2024;12:1338855. doi:10.3389/fped.2024.1338855

59. Li E, Smithson L, Khan M, et al. Effects of Perinatal Stroke on Executive Functioning and Mathematics Performance in Children. J Child Neurol. 2022;37(2):133–140. doi:10.1177/08830738211063683

60. Gullifer JW, Titone D. Characterizing the social diversity of bilingualism using language entropy. Biling Lang Cogn. 2020;23(2):283–294. doi:10.1017/S1366728919000026

61. Govaert P, Smith L, Dudink J. Diagnostic management of neonatal stroke. Semin Fetal Neonatal Med. 2009;14(5):323–328. doi:10.1016/j.siny.2009.07.007

62. Gacio S, Munoz Giacomelli F, Klein F. Presumed perinatal ischemic stroke. A review. Arch Argent Pediatr. 2015;113(5). doi:10.5546/aap.2015.eng.449

63. Ghotra SK, Johnson JA, Qiu W, Newton A, Rasmussen C, Yager JY. Age at stroke onset influences the clinical outcome and health-related quality of life in pediatric ischemic stroke survivors. Dev Med Child Neurol. 2015;57(11):1027–1034. doi:10.1111/dmcn.12870

64. Kirton A, DeVeber G, Pontigon AM, Macgregor D, Shroff M. Presumed perinatal ischemic stroke: Vascular classification predicts outcomes. Ann Neurol. 2008;63(4):436–443. doi:10.1002/ana.21334

65. Dennis M. Developmental plasticity in children: the role of biological risk, development, time, and reserve. J Commun Disord. 2000;33(4):321–332. doi:10.1016/S0021-9924(00)00028-9

66. Newport EL, Landau B, Seydell-Greenwald A, et al. Revisiting Lenneberg’s hypotheses about early developmental plasticity: Language organization after left-hemisphere perinatal stroke. Biolinguistics. 2017;11:407. doi:10.5964/bioling.9105

67. Lowe CJ, Cho I, Goldsmith SF, Morton JB. The Bilingual Advantage in Children’s Executive Functioning Is Not Related to Language Status: A Meta-Analytic Review. Psychol Sci. 2021;32(7). doi:10.1177/0956797621993108

68. Calvo A, Bialystok E. Independent effects of bilingualism and socioeconomic status on language ability and executive functioning. Cognition. 2014;130(3). doi:10.1016/j.cognition.2013.11.015

69. Meir N, Armon-Lotem S. Independent and combined effects of socioeconomic status (SES) and bilingualism on children’s vocabulary and verbal short-term memory. Front Psychol. 2017;8:1442. doi:10.3389/fpsyg.2017.01442

70. Brito NH, Noble KG. The independent and interacting effects of socioeconomic status and dual-language use on brain structure and cognition. Dev Sci. 2018;21(6). doi:10.1111/desc.12688

71. Peristeri E, Silleresi S, Tsimpli IM. Bilingualism effects on cognition in autistic children are not all-or-nothing: The role of socioeconomic status in intellectual skills in bilingual autistic children: Autism. Published online February 1, 2022. doi:10.1177/13623613221075097

72. De Lamo White C, Jin L. Evaluation of speech and language assessment approaches with bilingual children. Int J Lang Commun Disord. 2011;46(6):613–627. doi:10.1111/j.1460-6984.2011.00049.x

73. Freeman MR, Schroeder SR. Assessing Language Skills in Bilingual Children: Current Trends in Research and Practice. J Child Sci. 2022;12(1). doi:10.1055/s-0042-1743575

74. Fujii DEM. Developing a cultural context for conducting a neuropsychological evaluation with a culturally diverse client: the ECLECTIC framework*. Clin Neuropsychol. 2018;32(8). doi:10.1080/13854046.2018.1435826

